# Airborne contamination of COVID-19 in hospitals: a scoping review of the current evidence

**DOI:** 10.1101/2020.09.09.20191213

**Authors:** Gabriel Birgand, Nathan Peiffer-Smadja, Sandra Fournier, Solen Kerneis, François-Xavier Lescure, Jean-Christophe Lucet

**Author notes:** **Correspondence to**: Dr Gabriel Birgand, Address: 5 rue du Professeur Yves Boquien 44093 Nantes, France. E-mail address (G. Birgand).

## Abstract

**Introduction:** A controversy remains worldwide regarding the transmission routes of SARS-CoV-2 in hospital settings. We reviewed the current evidence on the air contamination with SARS-CoV-2 in hospital settings, and the factors associated to the contamination including the viral load and the particles size.

**Methods:** The MEDLINE, Embase, Web of Science databases were systematically interrogated for original English-language articles detailing COVID-19 air contamination in hospital settings between 1 December 2019 and 21 July 2020. This study was conducted in accordance with the PRISMA-ScR guidelines. The positivity rate of SARS-CoV-2 viral RNA and culture were described and compared according to the setting, clinical context, air ventilation system, and distance from patient. The SARS-CoV-2 RNA concentrations in copies per m^3^ of air were pooled and their distribution were described by hospital areas. Particle sizes and SARS-CoV-2 RNA concentrations in copies or TCID50 per m^3^ were analysed after categorization of sizes in < 1 µm, 1–4 µm, and > 4 µm.

**Results:** Among 2,034 records identified, 17 articles were included in the review. Overall, 27.5% (68/247) of air sampled from close patients’ environment were positive for SARS-CoV-2 RNA, without difference according to the setting (ICU: 27/97, 27.8%; non-ICU: 41/150, 27.3%; p = 0.93), the distance from patients (< 1 meter: 1/64, 1.5%; 1–5 meters: 4/67, 6%; p = 0.4). In other areas, the positivity rate was 23.8% (5/21) in toilets, 9.5% (20/221) in clinical areas, 12.4% (15/121) in staff areas, and 34.1% (14/41) in public areas. A total of 78 viral cultures were performed in three studies, and 3 (4%) were positive, all from close patients’ environment. The median SARS-CoV-2 RNA concentrations varied from 1.10^3^ copies per m^3^ (IQR: 0.4.10^3^-9.10^3^) in clinical areas to 9.7.10^3^ (5.1.10^3^-14.3.10^3^) in the air of toilets or bathrooms. The protective equipment removal and patients’ rooms had high concentrations/titre of SARS-CoV-2 with aerosol size distributions that showed peaks in the < 1 µm region, and staff offices in the > 4µm region.

**Conclusion:** In hospital, the air near and away from COVID-19 patients is frequently contaminated with SARSCoV-2 RNA, with however, rare proofs of their viability. High viral loads found in toilet/bathrooms, staff and public hallways suggests to carefully consider these areas.

## Introduction

A controversy remains worldwide regarding the transmission modes of SARS-CoV-2 virus. [1] At the emerging stage of the pandemic, many countries implemented a high-level precautions, including airborne and contact precautions, to prevent the spread from patients to healthcare professionals (HCP).[2] An emerging understanding of the SARS-CoV-2 transmission dynamics led to recommendations toward droplet precautions to care for hospitalized COVID-19 patients.[3] However, separating transmission dynamics into the dichotomy of droplet versus airborne transmission routes is probably simplistic. In some circumstances, large droplets may travel more than the commonly used 1.50 meters, and contaminate surfaces further away.[4]

The environment airflow may ease the spread of large particles.[5] The switch from airborne to droplet precautions, combined with a global shortage of facemasks and respirators, fed the controversy around respiratory protections for the prevention of transmission of SARS-CoV-2. [6,7] This generated a mistrust in personal protective equipment’s (PPE), especially surgical mask, and, in their ability to protect HCP from SARS-CoV-2 transmission. As WHO recently acknowledged that a possible airborne transmission could occur in crowded and closed environments in the community. The question arises whether similar transmission could occur in the hospital.[1] Viral contamination of the air surrounding COVID-19 patients and professionals in hospitals may have serious implications for outbreak control strategies.

We reviewed the current evidence on the air contamination with SARS-CoV-2 in hospital settings, the viral load, and the associated factors, to better assess the risk of cross transmission of COVID-19 among professionals and patients.

## Methods

### Search strategy

We performed a systematic search of MEDLINE via PubMed, Embase, Web of Science, on 21 July 2020 with the terms covering COVID-19, air contamination in hospital settings in articles published between 1 December 2019 and 21 July 2020 (see Supplement S1 for details of search strategies). Because of potential delays in indexing of databases, we also searched selected infectious disease journals (Supplementary Table S1). We also searched some preprint servers, including BioRxiv (https://www.biorxiv.org/). MedRxiv (https://www.medrxiv.org/); and reference lists of the identified articles to find reports of additional studies.

### Inclusion and exclusion criteria

We included all literature related to COVID-19 published in English between 1 January 2020 and 21 July 2020 without restrictions, including original articles, research letters and comments. We excluded experimental methods and studies performed in dental and primary care settings.

### Article selection and data extraction

Two reviewers screened all titles, abstracts and full texts independently and solved disagreements by consensus or consultation with a third reviewer. Then the following information were extracted: (i) the setting, (ii) the clinical context, (iii) the ventilation system, (iv) the number of air samples performed, (v) the sampling method, (vi) the location of sampler and distance from patients, (vi) the duration and air volume sampled, (vii) the method for SARS-CoV-2 search, (vii) the positivity rate, (viii) the viral load (SARS-CoV-2 RNA copies/m^3^), (ix) viral culture results. The details of the data collection are shown in Supplementary Table S2.

### Data analysis

We conducted a descriptive analysis of the characteristics of the included literature. We described the setting, patient clinical contexts, ventilation, air sampling and SARS-CoV-2 search methods, and the qualitative and quantitative results according to settings and the hospital area. We categorized the location of air sampling in five classes of hospital areas: close patient’s environment (i.e. patient’s room or bay), toilet/bathroom, clinical areas (workstation, anteroom/buffer room, corridors, other, in the clinical unit), staff areas (change room, staff room including office, meeting, dining rooms, others staff areas), public areas (hallway, other public areas indoor and outdoor). We also classified the setting in ICU and non-ICU, the clinical context in severe/critical vs mild/moderate/asymptomatic, the ventilation system in negative pressure vs natural/mechanical, and the distance from patient into ≤1, 1-5 and > 5 meters. The positivity rate viral RNA and the viral culture were described and compared according to categories using a chi2 test. The results of SARS-CoV-2 RNA concentrations in copies per m^3^ of air were pooled and their distribution were described by hospital areas. The Kruskall-Wallis test was used to compare the non-normal distributed RNA concentrations across hospital areas. Studies presenting the joined results of particle sizes and SARS-CoV-2 RNA concentrations in copies or TCID50 per m^3^ were analyzed after categorization of sizes in <1 µm, 1–4 µm, and >4 µm.

We conducted this scoping review in accordance with the PRISMA-ScR Checklist [16] (Supplementary Table S3).

## Results

### Search results

We identified 2,034 records, 280 of which were excluded as duplicates. Title and abstract screening were conducted for the remaining 1,364 articles, 1,225 of which were excluded because of being unrelated to the air contamination by SARS-CoV-2 in hospital settings. We retrieved the full text of the 139 remaining articles. After further screening, and supplementary searching of articles published or posted between 1 January 2020 and 21 July 2020, we identified an additional article and a total of 17 articles were included in the review (Figure 1).[3,8–23]

**Figure 1.**
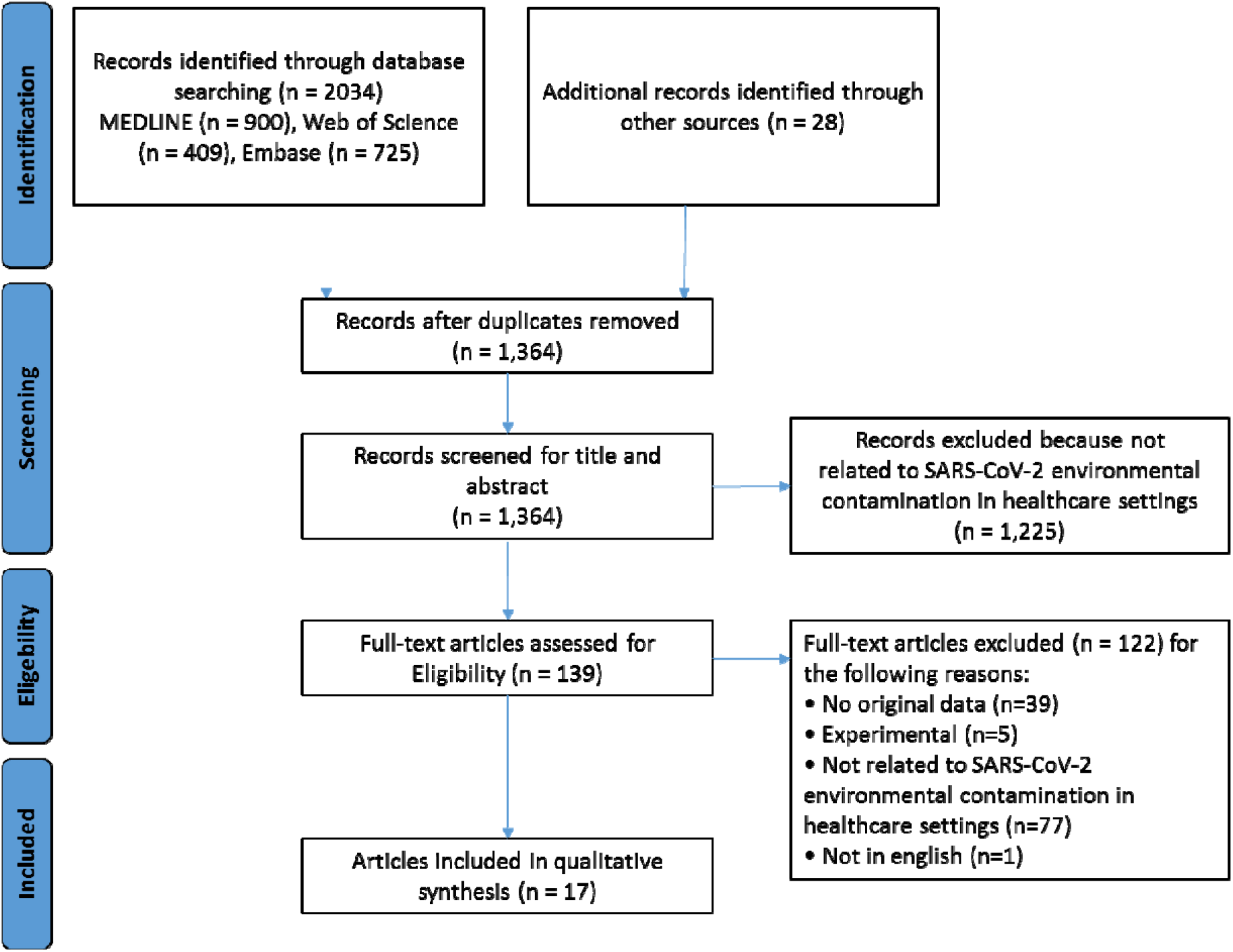
Flowchart of the search strategy.

### Characteristics of included articles/studies

Of the 17 included articles/studies, eight (55%) were from China. The remaining studies were from Hong Kong (n = 2), USA (n = 2), Singapore (n = 2), United Kingdom (n = 1), Italy (n = 1), Iran (n = 1). Of all included articles/studies, 13 (76%) were published in peer-reviewed journals and four (24%) were posted on preprint servers.

A total of 16 (94%) studies sampled the air in the close patients’ environment, 10 studies (59%) in clinical areas away from patients, 7 (41%) in staff areas, 5 (29%) in toilet/bathroom, and 5 (29%) in public area. (Table 1) The clinical context of patients hospitalized in the targeted areas was detailed in 11 studies among which six were performed in units hospitalizing severe/critical patients and seven mild/moderate/asymptomatic patients.

**Table 1.** Summary of included studies evaluating the air contamination with SARS-CoV-2 in the hospital environment.

| Ref | Setting | Clinical context | Location | Air ventilation | Distance from Pts, meters | Duration and air volume per sample | Microbiology | SARS-CoV-2 Viral load (RNA copies/liter) | SARS-CoV-2 Viral culture | Positivity, n/N (%) |
| --- | --- | --- | --- | --- | --- | --- | --- | --- | --- | --- |
| **ICU Patients environment** |  |  |  |  |  |  |  |  |  |  |
| [9] | ICU | Severe | Isolation rooms | Negative Pressure | ND | 5h to 7 days, 1500 – 50400L | ddPCR | 31, 113 | - | 2/3 (66) |
| [8] | ICU | Severe | Multiple beds room | Mechanical/Natural | 2 to 5 | 60 min/90L | RT-PCR, CT=38 | - | - | 0/9 (0) |
| [11] | ICU | Severe | Isolation rooms, bay room | 12 air supplies; 16 air discharges/h | ND | 30 min, 300L | RT-qPCR | Mean 3.8 near air outlet, 1.4 near Pts. | - | 13/32 (40.6), CT= 35.7, 44.4 |
| [22] | ICU | Critical | Multiple beds room | Not detailed | 1 | 4h, 1260L | RT-qPCR | - | - | 0/1 (0), CT= 41.5 |
| [10] | ICU | Severe | Multiple beds room (n=3) | 12 air supplies; 16 air discharges/h | 1 to 5 | 5h, 21600L | RT-PCR CT <37; >40 | - | - | 0/9 (0) |
| [19] | ICU | Not detailed | ND | Natural ventilation | 1 to 5 | 40 min; 600-16000L | RT-qPCR, CT=39.5 | NP | - | 0/5 (0) |
| [23] | ICU | 2 Pts intubated, 1 non intubated | Multiple beds room | Negative Pressure | ND | 40 min, 2000L | RT-PCR | Mean 22.7 | - | 12/12 (100) |
| [18] | ICU | Mild | Isolation rooms | Negative Pressure | 0.5 | 20-30min, 300-10000L | RT-PCR | - | - | 0/26 (0) |
| **Non-ICU Patients environment** |  |  |  |  |  |  |  |  |  |  |
| [9] | GW | Severe | 4 Single and 1 multiple bed room (n=2) | Negative Pressure | ND | 5h, 1500L | ddPCR | 0 | - | 0/2 (0) |
| [8] | GW | Severe | Multiple beds room (n= 2 to 9) | Mechanical/Natural | 2 to 5 | 60 min/90L | RT-PCR, CT=38 | - | - | 0/1 (0) |
| [3] | GW | Moderate/mild | Isolation rooms | 12 air exchanges per hour | <1 and 2-5 | 4h, 3600L | RT-PCR, CT=45 | Not determined | - | 0/18 (0) |
| [12] | IW | Mild | Isolation rooms | Negative Pressure | bedside table or desk | 15 min, 750L | RT-PCR, CT=45 |  | 0/10 | 10/18 (0) |
| [11] | GW | Mild | Single, bay room | 8 air supplies and 12 air discharges per hour | Near Pts, near air outlet | 30 min, 300L | RT-qPCR | 0.68 near air outlet | - | 2/16 (12.5) |
| [16] | GW | Mild/Asympto | Isolation rooms | 12 air changes per | 1 and 2.1 | 4h, 840L | RT-PCR | Mean 1.3 (Min | - | 4/10 (40) |
|  |  | matic |  | hour |  |  |  | 0.916-Max 2) |  |  |
| [22] | IW | Critical | ND | ND | 1 | 4h, 1260L | RT-qPCR | - | - | 1/1 (100)<br>CT=44.6 |
| [14] | IW | Severe/mild/asymptomatic | Isolation rooms | 12 air changes per hour, shelter | 0.1 | 20min, 1000L | RT-PCR | 0 | - | 0/6 (0) |
| [21] | IW | ND | Isolation rooms | Negative Pressure | 0.1 | 5min, 1000L | RT-PCR | 0 | - | 0/8 (0) |
| [17] | IW/GW | ND | ND | Not detailed | ND | ND, 1000L | RT-qPCR<br>CT<40.4 | 1.17 (0.16-7.04) | 0 | 6/12 (50) |
| [10] | IW | Severe | Multiple beds room (n=3) | 8 air supplies; 12 air discharges/h | 1 to 5 | 5h, 21600L | RT-PCR<br>CT <37; >40 | - | - | 0/18 (0) |
| [15] | IW | Mild/Asymptomatic | Isolation rooms | 12 air exchanges per hour | 0.6 | 15 min, 1500L | RT-PCR,<br>CT<35 | - | - | 0/6 (0) |
| [19] | GW | ND | Isolation rooms | Natural | 1 to 5 | 40 min, 600-16000L | RT-PCR,<br>CT <39.5 | - | - | 0/16 (0) |
| [13] | ND | ND | Isolation rooms | Not detailed | ND | 30 min, ND | RT-qPCR | 2.41 TCID50/cm3 of Air | 3/18 | 18/18 (100) |
| **Toilet/Bathroom** |  |  |  |  |  |  |  |  |  |  |
| [9] | Non-ICU | - | Patient mobile toilet room | No Ventilation | - | 20h, 6000L | ddPCR | 1 | - | 1/1 (100) |
| [3] | GW | - | Isolation rooms | ND | - | 8h, 2400L | RT-PCR,<br>CT=45 | - | - | 0/6 (0) |
| [22] | Isolation ward | - | Patient bathroom | ND | - | 4h, 1680L | quantitative RT-PCR | - | - | 2/2 (100)<br>CT=35.6, 35.5 |
| [17] | Cohort ward | - | Outside the patients' bay, in the ward | ND | - | ND, 1000L | RT-qPCR<br>CT<40.4 | 1/2, 0.464 | Negative | 1/2 (50) |
| [19] | Fever clinic | - | In the ward | Natural | - | 40, 16000L | RT-PCR, CT <39.5 | - | - | 0/3 (0) |
| [18] | 4 in ICU | - | Patient bathroom | 4 with negative pressure | - | 20-30min, 420-10000L | RT-PCR | - | - | 1/7 (14) |
| **Clinical areas** |  |  |  |  |  |  |  |  |  |  |
| [9] | Non-ICU | ND | Workstation | Natural | ND | 320-1200min, 1600-6000L | ddPCR | 0, 1, 1, 5, 9 | - | 4/5 (80) |
| [3] | ICU, Non-ICU | ND | Corridor, anteroom | - | ND | 15min/3000L, 480min/2400L | RT-PCR,<br>CT=45 | - | - | 0/12 (0) |
| [12] | ND | Mild | Floor adjacent to rooms | - | > 6ft | 15min/750L | RT-PCR,<br>CT=45 | 2.58, 3.76 | 0/3 | 2/3 (66) |
| [11] | ICU, GW | 1 severe, ND | Near office, pharmacy, nurse station, corridor,<br>buffer room | ND | ND | 30min, 27000-72000L | RT-qPCR | 0.54 | - | 1/34 (3) |
| [20] | 1 ICU | Ward, and<br>various clinical<br>rooms | ND | ND | ND | 30min/ND | RT-PCR,<br>CT=43 | - | - | 0/69 (0) |
| [17] | ICU,<br>Non-ICU | ND | Nurse station,<br>Ward,<br>Ambulatory<br>waiting, Resus<br>bay, CPAP unit | ND | ND | ND, 1000L | RT-qPCR<br>CT<40.4 | 0.404,<br>0.035,1.922,<br>0.031 | 0/10 | 4/10 (40) |
| [10] | ICU,<br>Non-ICU | Severe | Corridor, clinic,<br>buffer room | 1 with Negative<br>pressure | ND | 250min,<br>21600L | RT-PCR<br>CT <37; >40 | - | - | 0/45 (0) |
| [19] | ND | ND | Corridor, pre-<br>room | Natural | ND | 40min, 16000L | RT-PCR,<br>CT <39.5 | - | - | 0/18 (0) |
| [23] | ND | ND | Corridor | ND | ND | 40min, 16000L | RT-PCR | - | - | 8/8 (100),<br>CT=31.1 |
| [18] | IW | 3 mild, ND for<br>others | Ward, corridor,<br>nurse station,<br>storage room | 3 negative pressure,<br>ND for others | ND | 20-30, 300-10000 | RT-PCR | - | - | 1/12 (8),<br>CT=37.8 |
| **Staff areas** |  |  |  |  |  |  |  |  |  |  |
| [9] | ICU | - | Change, meeting,<br>dinning,<br>warehouse | Natural/mechanical,<br>small air purifier | - | 300-1200min/1500-6000L | ddPCR | Mean: 13.8 | - | 10/13 (77) |
| [12] | Isolation<br>ward | - | Personal air<br>sample | - | ND | - | RT-PCR,<br>CT=45 | Mean: 20.037 | - | 4/4 (100) |
| [11] | ICU/GW | - | Dressing rooms | - | ND | 30min/45000-108000L | RT-qPCR | - | - | 0/36 (0) |
| [17] | ND | - | Staff room,<br>doffing room | - | ND | ND/1000L | RT-qPCR<br>CT<40.4 | 0.249 | 0/4 | 1/4 |
| [10] | ICU/IW | - | Conference room,<br>clean zone | 2/3 with negative<br>pressure | ND | 270-540min/21600-43200L | RT-PCR<br>CT <37; >40 | - | - | 0/45 |
| [19] | ND | - | Waste storage | Natural | ND | 40min/16000L | RT-PCR, CT<br><39.5 | - | - | 0/2 |
| [23] | ND | - | Dressing/undressi<br>ng, lockers | - | ND | 40min/8000-14000L | RT-PCR | Not determined | - | 0/17 |
| **Public areas** |  |  |  |  |  |  |  |  |  |  |
| [9] | - | - | Pharmacy, hall,<br>office, store,<br>supermarket | Mechanical, natural,<br>outdoor | - | 300-1000min,<br>1500-5000L | ddPCR | 3, 7, 11, 3 | - | 4/11 |
| [12] | - | - | Hallway | ND | - | ND | RT-PCR,<br>CT=45 | 0.979 to 8.688 | 0/12 | 8/12 |
| [20] | - | - | Public area | ND | - | 30min/ND | RT-PCR,<br>CT=43 | - | - | 0/6 |
| [17] | - | - | Main entrance,<br>toilet entrance, lift<br>area | ND | - | ND, 1000L | RT-qPCR<br>CT<40.4 | 1.574,<br>1.545 | 0/3 | 2/3 |
| [10]i | - | - | Public area | ND | - | 270min,<br>21600L | RT-PCR<br>CT <37; >40 | - | - | 0/9 |

Abbreviations: ICU, intensive care unit ; IW, isolation ward; ND, not detailed; ddPCR, droplet-digital polymerase chain reaction; RT-PCR, reverse transcriptase polymerase chain reaction; RT-qPCR, quantitative reverse transcriptase polymerase chain reaction; CT, cycle threshold; Pts, patients; GW, general wards; IR, isolation room; ft, feet.

A median of 35 air samples were collected per study varying from four to 126 samples. In the close patients’ environment, a median of 12 air samples (Min: 2; Max: 42) was performed, 2.5 (1–7) in toilets/bathrooms, 12 (3–72) in patients’ environment, 13 (2–45) in staff areas, and 9 (3–12) in public areas. Fourteen studies sampled the air from non-ICU patient’s rooms, and nine in ICU. Overall, 89 samples performed in patients’ room with negative pressure and 31 with natural/mechanical ventilation. When pooling the 12 studies detailing the distance from patient, a total of 49 samples were performed ≤1 meter from patients and 42 from one to five meters.

All included studies used RT-PCR to identify SARS-CoV-2 RNA, with a quantification of RNA copies per m^3^ or liters in 8 (47%) studies. One study used a droplet digital RT-PCR method. The viral culture was performed in three (18%) studies. Three studies assessed the particle size in parallel to RNA concentration or viral titer.[9,13,16]

### RT-PCR and culture results categorized by hospital areas

A total of 646 air samples were performed across the 17 studies reviewed, including 247 (38%) in close patient’s environment, 221 (34%) in clinical areas, 121 (19%) in staff area, 41 (6%) in public areas, and 21 (3%) in toilet/bathroom. (Table 2) Overall, 27.5% (68/247) of air sampled from close patients’ environment were positive for SARS-CoV-2 RNA. Among the 97 samples performed in the environment of ICU patients, 27 (27.8%) were positive, vs 41 among 150 (27.3%) in non-ICU rooms. The air RNA positivity rate was 23.4% (43/184) in room with negative pressure, and no sample out of 31 were positive in natural/mechanical ventilated rooms. In toilets/bathroom, 5/21 (23.8%) samples were positive. In clinical areas, the overall positivity rate was 9.5% (20/221) varying from 0% (0/44) in anteroom/buffer room, to 27.2% (6/22) at workstations (p< 0.01). In staff areas, 12.4% (15/121) of samples were positive, with 19.2% of positivity in staff meeting room (5/26), vs 4% (2/50) in donning/doffing room and 17.8% (8/45) in other types of staff rooms (p = 0.06). Overall, 34% (14/41) of samples in public areas were positives with 60% (9/15) in hallways, 11% (2/18) in other indoors areas and 36% (3/8) in outdoors public areas (p = 0.01). A total of 78 viral cultures were performed across three studies (39 air samples from close patient’s environment, two in toilets/bathroom, 14 in patients area, eight in staff areas, 15 in public areas). Three samples were positive in one study from the close patients’ environment (3/39, 7.7%) in a non-ICU setting.

**Table 2.** Description of RT-PCR and culture results categorized by hospital areas.

|  | SARS-CoV-2 Viral RNA |  |  | SARS-CoV-2 Viral culture |  |
| --- | --- | --- | --- | --- | --- |
|  | n/N | Positivity, % | p value | n/N | Positivity, % |
| **Patients environment** | 68/247 | 27.5 |  | 3/39 | 7.7 |
| Ward |  |  |  |  |  |
| ICU | 27/97 | 27.8 | 0.93 | - | - |
| Non-ICU | 41/150 | 27.3 |  | 3/39 | 7.7 |
| Ventilation |  |  |  |  |  |
| Negative pressure | 43/184 | 23.4 | 0.03 | 0/10 | 0 |
| Mechanical/natural | 0/31 | 0 |  | - | - |
| Distance from patient |  |  |  |  |  |
| ≤1 meter | 1/64 | 0 | 0.19 | - | - |
| 1-5 meter | 4/67 | 6 |  | - | - |
| Clinical context |  |  |  |  |  |
| Severe/critical | 16/77 | 20.8 | 0.43 | - | - |
| Other (mild, moderate, asymptomatic) | 16/99 | 16.1 |  | 0/10 | 0 |
| **Patients' toilet/bathroom** | 5/21 | 23.8 |  | 0/2 | 0 |
| **Clinical areas** | 20/221 | 9.5 |  | 0/14 | 0 |
| Corridor | 9/47 | 19.1 | <0.01 | - | - |
| Workstation | 6/22 | 27.2 |  | 0/5 | 0 |
| Anteroom/Buffer room | 0/44 | 0 |  | - | - |
| Others | 5/103 | 4.8 |  | 0/9 | 0 |
| **Staff areas** | 15/121 | 12.4 |  | 0/8 | 0 |
| Donning/doffing room | 2/50 | 4 | 0.06 | 0/1 | 0 |
| Meeting/staff room | 5/26 | 19.2 |  | 0/3 | 0 |
| Others | 8/45 | 17.8 |  | 0/4 | 0 |
| **Public areas** | 14/41 | 34.1 |  | 0/15 | 0 |
| Hall/Hallway | 9/15 | 60 | 0.01 | 0/14 | 0 |
| Others indoor | 2/18 | 11.1 |  | 0/1 | 0 |
| Hospital Outdoors | 3/8 | 37.5 |  | - | - |

### SARS-CoV-2 RNA concentrations in copies per m^3^ of air according to hospital areas

Among studies with SARS-CoV-2 positive air samples which performed a quantitative RT-PCR, the median RNA concentrations varied from 1.10^3^ copies per m^3^ (IQR: 0.4.10^3^-9.10^3^) in clinical areas to 9.7.10^3^ (5.1.10^3^-14.3.10^3^) in the air of toilets/bathrooms. (Figure 2) The median concentration described in close patients’ environment was 3.3.10^3^ (1.10^3^-2.7.10^3^) copies per m^3^ (p< 0.01). Among the three studies which assessed the particle size in air sampled in parallel with the viral load, one found a RNA concentration of 2.10^3^copies/m^3^ for size > 4 µm and 1.3.10^3^ for 1–4 µm particles in one patients’ room, and 927 and 916 copies/m^3^ respectively in a second room, both at a distance of 1 to 2.1 meters from patients. (Figure 3) A second study found 40 and 12.10^3^ copies/m^3^ for < 1-4 µm particles in two PPE removal rooms. A concentration of 7.10^3^ copies/m^3^ was found for < 1 µm particles and 13.10^3^ copies/m^3^ for 1-4 µm particles in medical staff office. For the third study which performed viral culture with air samples from six different patients’ room, the median viral concentration was 4.8 (3.3–5.8) TCID50/m^3^ for < 1 µm particles, 4.27 (2.96–5.48) for 1–4 µm particles, and 1.82 (1.6–2.55) for > 4 µm particles.

**Figure 2.**
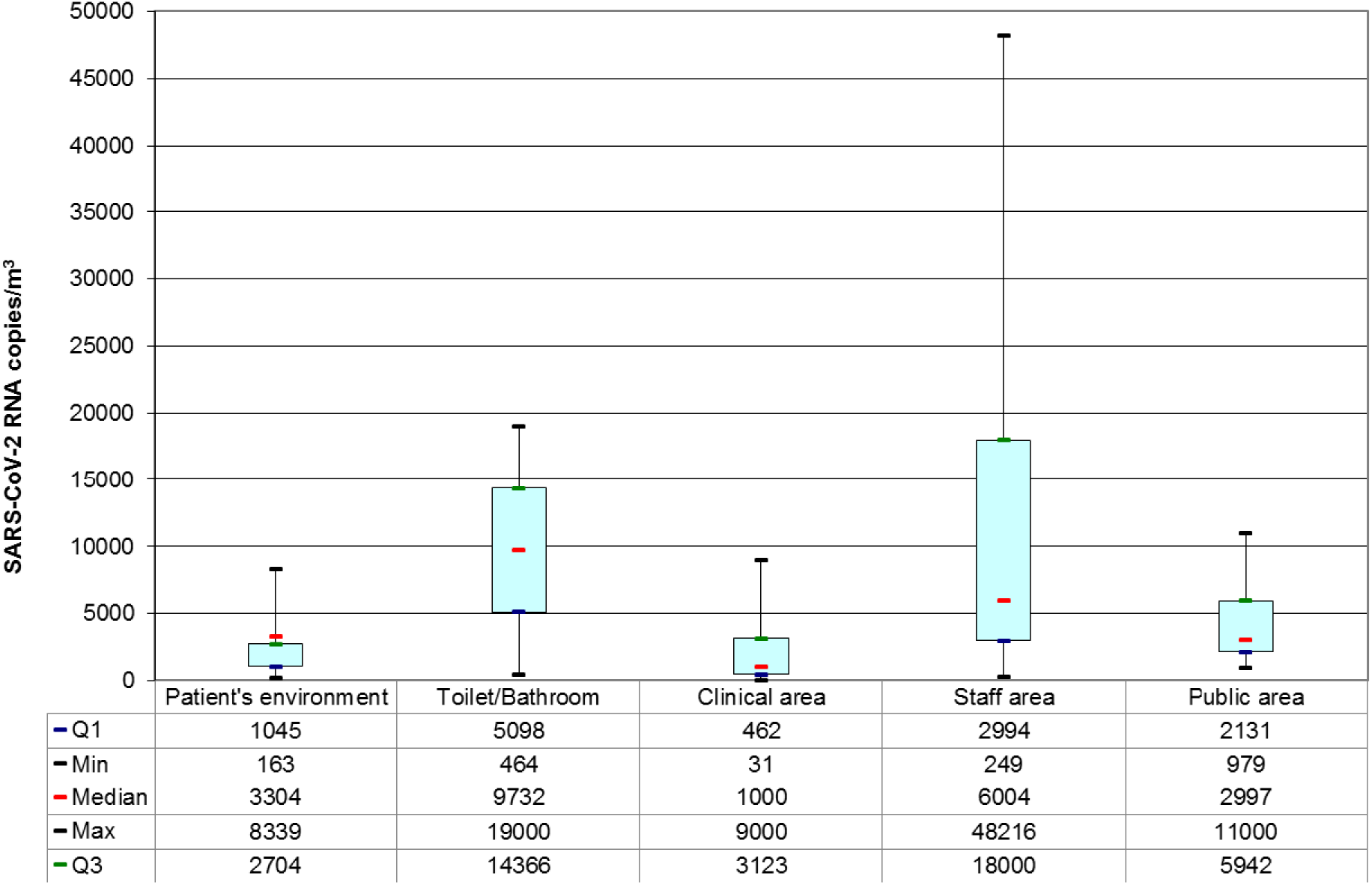
Distribution of the pooled SARS-CoV-2 RNA concentrations in copies per m^3^ of air according to hospital areas.

**Figure 3.**
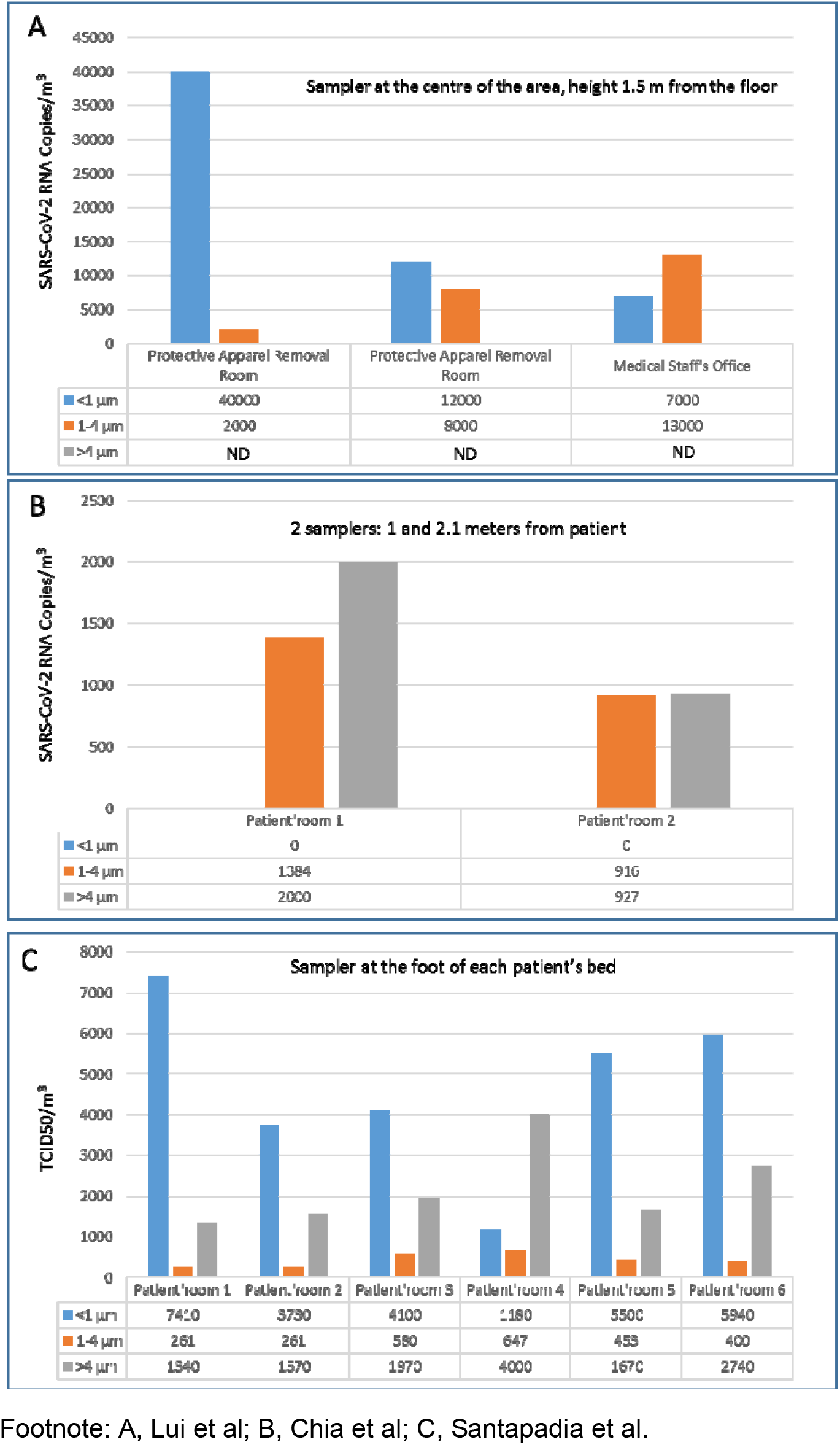
Concentration of airborne SARS-CoV-2 in different aerosol size.

## Discussion

This scoping review of the literature suggests the air near hospitalized patients, but also away from patients’ environment including toilet/bathrooms, staff and public areas, may carry viral RNA. However, the infectivity of the virus assessed by viral culture was only reported by one study in non-ICU patients’ room. The protective equipment removal and patients’ rooms had high concentrations/titre of SARS-CoV-2 with aerosol size distributions that showed peaks in the < 1 µm region, and staff offices in the > 4 µm region.

The results of positivity rate in ICU and non-ICU patients’ environment was highly heterogeneous, and appeared equal when pooling the results. In ICU, 5/8 studies did not find SARS-CoV-2 RNA, whereas the remaining did with 40.4 to 100% of positive samples. In non-ICU patients’ environment, 9/14 did not find SARS-CoV-2 RNA, and five studies found viral RNA present from 12.5% to 100% of samples. This heterogeneity may be explained either by a different case-mix, or by a difference in methods used for air sampling. The level of severity of patients’ infections was not associated with an increased air contamination. Several studies suggested higher viral loads might be associated with severe clinical outcomes.[24,25] However, the correlation between clinical conditions and the air contamination may be more complex. The potential opportunistic airborne contamination occurring during aerosol generating procedures (AGP), and the ventilation at the time of sampling may influence the results. All these factors were poorly detailed in the articles analyzed. The sampling method, including the sampler used, its position in the clinical unit and from patients, the duration of sampling, the volume sampled, and condition for transfer to the laboratory were highly variable across studies. The volume of a single room is approximately 40 m^3^. However, most samples volume were less than 10 m^3^, at various airflow rates, for a duration less or equal to one hour, potentially not reflecting the reality of the air contamination. The climatic conditions (e.g. temperature and hygrometry), were poorly detailed in studies reviewed but may influence the capacity for viral particles to persist in the air.[26] The methods for RNA detection varied, especially the cycle threshold (Ct) for PCR positivity also varied from 37 to 45. The RT-PCR Ct values correlate strongly with cultivable virus. A recent article showed the probability of culturing virus declines to 8% in samples with Ct > 35.[27] Only one study described a positive viral culture on samples with SARS-CoV-2 RNA on RT-PCR, suggesting that most samples do not contain enough infectious virus. Most sampling methods affect viral infectivity, which may partly explain these results.[28] The future studies should consider these points for a better accuracy and comparability of the data.

The concentration of SARS-CoV-2 RNA in aerosols detected in isolation wards and ventilated patients was very low. However, a higher concentration of viral RNA was found in patients’ toilets, public areas and in some medical staff areas. The finding of high concentration in staff room (meeting, dining room) is consistent with the possible cross-transmission of COVID-19 among HCP during breaks. During these periods, facemasks are frequently removed in small areas without ventilation. Toilets and staff rooms are often small, and poorly ventilated. The presence of SARSCoV-2 RNA in stools has been described in several studies. [29,30] The toilet flush may lead to the aerosolization of RNA in small and non-ventilated toilets or bathrooms. Public areas are often crowded in such epidemic context with both a high patient flow and high incidence of COVID-19. These factors have to be considered to control the transmission of COVID-19 between non-masked HCPs in hospital setting, especially staff rooms and lockers.

Only three studies assessed the size of particle found when searching for SARS-CoV-2. Regarding the aerosols in the submicrometre region, observed in the PPE removal and patients’ rooms, authors hypothesized the resuspension of virus-laden aerosols from the surface of the protective apparel worn by medical staff while they are removing the equipment. The submicrometre virus-laden aerosols may originally come from the direct deposition of respiratory droplets or airborne SARS-CoV-2 from a patient onto the protective apparel. On the other hand, floor-deposited SARSCoV-2 is possibly the source of > 4 µm virus-laden aerosols and was carried across different areas by medical staff.

The findings of this scoping review suggest COVID-19 to be an opportunistic airborne infection. These results are consistent with the cumulated knowledge on other respiratory viruses. SARSCoV-1 is commonly recognized to be mainly transmitted through large droplets, and requiring particular conditions to be airborne transmitted, such as AGP.[31] [32] For other respiratory viruses, a recent review described the frequent presence of nucleic material (RNA or DNA) in the air around infected patients (influenza, RSV, adenovirus, rhinovirus, and others coronavirus), but rarely the presence of viable viruses.[33] The current available evidence on hospital air contamination by SARS-CoV-2 leans toward the effectiveness of surgical facemask in most circumstances to prevent cross-transmission of COVID-19 in hospital settings. In contrast, AGP on the respiratory tract require wearing a respirator (N95 or FFP2), in order to prevent transmissions and protect HCPs.[5,34] A randomized clinical trial comparing the surgical facemask with respirator may provide important information for the recommending or not respiratory protection for HCP in addition for AGP. Assessing the SARS-CoV-2 RNA and viable virus contamination of surgical facemasks and respirators worn by HCP according to a panel of procedures with COVID-19 patients would provide information on the exposition in routine practice.

This study also has some limitations. First, the context (location, ventilation, distance, clinical context) were infrequently detailed in studies. As explained above, this potentially affected the comparability of data, and the reliability of pooled data analysis. Misclassification may have occurred when variables were categorized without enough details. This issue was avoided by performing the categorization only when data were available. Second, for a better clarity of the analysis, we did not include contamination of surfaces. However, air and surface contamination are potentially correlated, and may ease the understanding of resuspension. Finally, we included article not validated through a peer review process.

## Conclusion

In hospitals, the air around COVID-19 patients is frequently contaminated with SARS-CoV-2 RNA, but rarely with viable viruses. The available data suggests COVID-19 to be an opportunistic airborne infection. High viral loads found in toilet/bathrooms, staff and public hallways argue for a careful consideration of these areas for the prevention of the COVID-19 transmission. However, the presence of viable virus should be primarily considered, as the required link for the potential of cross-transmission.

## Data Availability

All data are available

## Declarations

### Ethics approval and consent to participate

not applicable

### Consent for publication

not applicable

### Availability of data and material

Data sharing not applicable to this article as no datasets were generated or analysed during the current study

### Funding

The research was funded by the National Institute for Health Research Health Protection Research Unit (NIHR HPRU) in Healthcare Associated Infection and Antimicrobial Resistance at Imperial College London in partnership with Public Health England (PHE). The views expressed are those of the author(s) and not necessarily those of the NHS, the NIHR, the Department of Health or Public Health England. GB has received an Early Career Research Fellowship from the Antimicrobial Research Collaborative at Imperial College London, and acknowledges the support of the Welcome trust. RA is supported by a NIHR Fellowship in knowledge mobilisation. The support of ESRC as part of the Antimicrobial Cross Council initiative supported by the seven UK research councils, and also the support of the Global Challenges Research Fund, is gratefully acknowledged.

### Competing Interests

The authors declare that they have no competing interests.

## Acknowledgements

We thanks Marta Castrica and Laura Menchetti for providing their data.

## Notes

### Competing Interest Statement

None

## REFERENCES

1. World Health Organisation. Transmission of SARS-CoV-2: implications for infection prevention precautions. 2020; Available at: https://www.who.int/newsroom/commentaries/detail/transmission-of-sars-cov-2-implications-for-infection-preventionprecautions.

2. Birgand G, Mutters NT, Otter JA, et al. Analysis of national and international guidelines on respiratory protection equipment for COVID-19 in healthcare settings. 2020;

3. Ong SWX, Tan YK, Chia PY, et al. Air, Surface Environmental, and Personal Protective Equipment Contamination by Severe Acute Respiratory Syndrome Coronavirus 2 (SARS-CoV-2) From a Symptomatic Patient. JAMA 2020;

4. Bourouiba L. Turbulent Gas Clouds and Respiratory Pathogen Emissions: Potential Implications for Reducing Transmission of COVID-19. JAMA 2020;

5. Seto WH. Airborne transmission and precautions: facts and myths. J Hosp Infect 2015; 89: 225–228.

6. Morawska L, Tang JW, Bahnfleth W, et al. How can airborne transmission of COVID-19 indoors be minimised? Environ Int 2020; 142: 105832.

7. Chagla Z, Hota S, Khan S, Mertz D, International Hospital and Community Epidemiology Group. Airborne Transmission of COVID-19. Clin Infect Dis 2020;ciaa1118.

8. Faridi S, Niazi S, Sadeghi K, et al. A field indoor air measurement of SARS-CoV-2 in the patient rooms of the largest hospital in Iran. Sci Total Environ 2020; 725: 138401.

9. Liu Y, Ning Z, Chen Y, et al. Aerodynamic analysis of SARS-CoV-2 in two Wuhan hospitals. Nature 2020;

10. Li YH, Fan YZ, Jiang L, Wang HB. Aerosol and environmental surface monitoring for SARSCoV-2 RNA in a designated hospital for severe COVID-19 patients. Epidemiol Infect 2020;1–14.

11. Guo Z-D, Wang Z-Y, Zhang S-F, et al. Aerosol and Surface Distribution of Severe Acute Respiratory Syndrome Coronavirus 2 in Hospital Wards, Wuhan, China, 2020. Emerg Infect Dis 2020; 26.

12. Santarpia JL, Rivera DN, Herrera V, et al. Aerosol and Surface Transmission Potential of SARS-CoV-2. Infectious Diseases (except HIV/AIDS), 2020. Available at: http://medrxiv.org/lookup/doi/10.1101/2020.03.23.20039446. Accessed 16 July 2020.

13. Santarpia JL, Herrera VL, Rivera DN, et al. The Infectious Nature of Patient-Generated SARSCoV-2 Aerosol. Infectious Diseases (except HIV/AIDS), 2020. Available at: http://medrxiv.org/lookup/doi/10.1101/2020.07.13.20041632. Accessed 22 July 2020.

14. Cheng VC-C, Wong S-C, Chan VW-M, et al. Air and environmental sampling for SARS-CoV-2 around hospitalized patients with coronavirus disease 2019 (COVID-19). Infect Control Hosp Epidemiol 2020;1–8.

15. Wei L., Lin J., Duan X., et al. Asymptomatic COVID-19 Patients Can Contaminate Their Surroundings: an Environment Sampling Study. mSphere 2020; 5.

16. Chia PY, Coleman KK, Tan YK, et al. Detection of Air and Surface Contamination by Severe Acute Respiratory Syndrome 2 Coronavirus 2 (SARS-CoV-2) in Hospital Rooms of Infected Patients. medRxiv. 2020; Available at: https://doi.org/10.1101/2020.03.29.20046557.

17. Zhou J, Otter JA, Price JR, et al. Investigating SARS-CoV-2 surface and air contamination in an acute healthcare setting during the peak of the COVID-19 pandemic in London. Clin Infect Dis Off Publ Infect Dis Soc Am 2020;

18. Ding Z, Qian H, Xu B, et al. Toilets dominate environmental detection of SARS-CoV-2 virus in a hospital. Infectious Diseases (except HIV/AIDS), 2020. Available at: http://medrxiv.org/lookup/doi/10.1101/2020.04.03.20052175. Accessed 16 July 2020.

19. Zhou L, Yao M, Zhang X, et al. Detection of SARS-CoV-2 in Exhaled Breath from COVID-19 Patients Ready for Hospital Discharge. Public and Global Health, 2020. Available at: http://medrxiv.org/lookup/doi/10.1101/2020.05.31.20115196. Accessed 16 July 2020.

20. Wu S, Wang Y, Jin X, Tian J, Liu J, Mao Y. Environmental contamination by SARS-CoV-2 in a designated hospital for coronavirus disease 2019. Am J Infect Control 2020;

21. Cheng VCC, Wong S-C, Chen JHK, et al. Escalating infection control response to the rapidly evolving epidemiology of the coronavirus disease 2019 (COVID-19) due to SARS-CoV-2 in Hong Kong. Infect Control Hosp Epidemiol 2020; 41: 493–498.

22. Lei H, Ye F, Liu X, et al. SARS-CoV-2 environmental contamination associated with persistently infected. Influenza Other Respir Viruses 2020;

23. Razzini K, Castrica M, Menchetti L, et al. SARS-CoV-2 RNA detection in the air and on surfaces in the COVID-19 ward of a hospital in Milan, Italy. Sci Total Environ 2020; 742: 140540.

24. Liu Y, Yan L-M, Wan L, et al. Viral dynamics in mild and severe cases of COVID-19. Lancet Infect Dis 2020; 20: 656–657.

25. Wolfel R., Corman V.M., Guggemos W., et al. Virological assessment of hospitalized patients with COVID-2019. Nature 2020; 581: 465–469.

26. Tang S, Mao Y, Jones RM, et al. Aerosol transmission of SARS-CoV-2? Evidence, prevention and control. Environ Int 2020; 144: 106039.

27. Singanayagam A, Patel M, Charlett A, et al. Duration of infectiousness and correlation with RTPCR cycle threshold values in cases of COVID-19, England, January to May 2020. eurosurveillance 2020; 25. Available at: https://www.eurosurveillance.org/content/10.2807/1560-7917.ES.2020.25.32.2001483. Accessed 31 August 2020.

28. Verreault D, Moineau S, Duchaine C. Methods for sampling of airborne viruses. Microbiol Mol Biol Rev MMBR 2008; 72: 413–444.

29. Wang W, Xu Y, Gao R, et al. Detection of SARS-CoV-2 in Different Types of Clinical Specimens. jama 2020; Available at: https://jamanetwork.com/journals/jama/fullarticle/2762997. Accessed 28 August 2020.

30. Wu Y, Guo C, Tang L, et al. Prolonged presence of SARS-CoV-2 viral RNA in faecal samples. Lancet Gastroenterol Hepatol 2020; 5: 434–435.

31. Roy CJ, Milton DK. Airborne transmission of communicable infection--the elusive pathway. N Engl J Med 2004; 350: 1710–1712.

32. Booth TF, Kournikakis B, Bastien N, et al. Detection of airborne severe acute respiratory syndrome (SARS) coronavirus and environmental contamination in SARS outbreak units. J Infect Dis 2005; 191: 1472–1477.

33. Shiu EYC, Leung NHL, Cowling BJ. Controversy around airborne versus droplet transmission of respiratory viruses: implication for infection prevention. Curr Opin Infect Dis 2019; 32: 372–379.

34. Seto WH, Tsang D, Yung RWH, et al. Effectiveness of precautions against droplets and contact in prevention of nosocomial transmission of severe acute respiratory syndrome (SARS). Lancet Lond Engl 2003; 361: 1519–1520.

